# Using a genetic risk score to estimate the earliest age of Alzheimer’s disease-related physiologic change in Body Mass Index

**DOI:** 10.1101/19013441

**Authors:** Willa D. Brenowitz, Scott C. Zimmerman, Teresa J. Filshtein, Kristine Yaffe, Stefan Walter, Thomas J. Hoffmann, Eric Jorgenson, Rachel A. Whitmer, M. Maria Glymour

## Abstract

**Objectives:** Weight loss is common in the years before an Alzheimer’s disease (AD) diagnosis, likely due to changes in appetite and diet. The age at which this change in body mass index (BMI) emerges is unclear but may point to the earliest manifestations of AD, timing that may be important for identifying windows of intervention or risk reduction. We examined the association between AD genetic risk and cross-sectional BMI across adults in mid-to late-life as an innovative approach to determine the age at which BMI changes and may indicate preclinical AD.

**Design:** Observational study

**Setting:** UK Biobank

**Participants:** 407,386 UK Biobank non-demented participants aged 39-70 with Caucasian genetic ancestry enrolled 2007-2010.

**Main Outcome Measures:** BMI (kg/m^2^) was constructed from height and weight measured during the initial visit. A genetic risk score for AD (AD-GRS) was calculated as a weighted sum of 23 genetic variants previously confirmed to be genome-wide significant predictors of AD (Z-scored). We evaluated whether the association of AD-GRS with BMI differed by age using linear regression with adjustment for sex and genetic ancestry, stratified by age grouping (40-60, 61+). We calculated the earliest age at which high AD-GRS predicted divergence in BMI compared to normal age-related BMI trends with linear and quadratic terms for age and interactions with AD-GRS.

**Results:** In 39-49 year olds, AD-GRS was not significantly associated with lower BMI (0.00 kg/m^2^ per SD in AD-GRS; 95%CI: -0.03,0.03). In 50-59 year olds AD-GRS was associated with lower BMI (-0.03 kg/m^2^ per 1 SD in AD-GRS; 95%CI:-0.06,-0.01) and this association was stronger in 60-70 year olds (-0.09 kg/m^2^ per 1 SD in AD-GRS; 95%CI:-0.12,-0.07). Model-based BMI age-curves for people with high versus low AD-GRS scores began to diverge after age 47.

**Interpretation:** Genetic factors that increase AD risk begin to predict lower BMI in adults by age 50, with greater effect later in older ages. Weight loss may manifest as an early pathophysiologic change associated with AD.

## INTRODUCTION

Given the long, insidious development of dementia and the subtle nature of early cognitive changes, it remains unclear at what age the earliest manifestations of Alzheimer’s disease (AD) emerge. Growing evidence of brain changes occurring decades prior to dementia diagnosis may help explain the failure of recent AD clinical trials, which enroll older adults with cognitive impairment.^1–3^ Identifying when the earliest indicators of AD occur would help to identify timing targets for when preventive interventions may be more effective. Weight loss may be an early indicator of incipient AD.^4–7^ Neurodegenerative changes,^8,9^ changes in metabolism, appetite, and nutrition^10^ or other metabolic factors may impact Body Mass Index (BMI) prior to clinical dementia onset. The age at which weight loss emerges in patients who continue on to develop AD is unclear but may point to the earliest manifestations of AD.

Identifying the earliest age of weight loss is challenging because higher midlife BMI may increase the risk of AD.^11–14^ Thus, the earliest weight loss resulting from developing AD may not be detectable when comparing earlier BMI values of people subsequently diagnosed with AD. To evaluate the age when AD-related weight loss begins, we adopt an approach using the genetic risk of AD. Genetic risk is determined at conception, prior to disease onset, but identifies individuals who are at high risk of developing AD in the future.^15,16^ Thus, any association between AD-related genetic variants and BMI cannot be attributed to the influence of BMI on AD, but instead would suggest that a biological process associated with the AD-GRS modifies BMI, assuming there is no pleiotropy of genetic risk for AD and BMI.^17^

The objective of the current study was to take advantage of known genetic variation in late-onset AD risk^15^ to identify the earliest age at which genetic risk for AD was associated with lower BMI (as a proxy for weight loss). Our approach parallels the Dominantly Inherited Alzheimer Network (DIAN) study, which assessed BMI in “preclinical AD” by comparing the point at which BMI curves began to diverge between autosomal dominant AD mutation carriers and non-carriers. In DIAN, mutation carriers had significantly lower BMI than non-carriers 11.2 years prior to expected symptom onset, and BMI curves began to diverge 17.8 years before expected symptom onset.^7^ The DIAN study focuses on early-onset AD due to specific genetic mutations, but the majority of AD cases are late-onset or sporadic AD which differs in etiology. To our knowledge, no prior study has used this approach to examine BMI trajectories associated with risk of late-onset AD. In this study, we evaluated the association between late-onset AD genetic risk and BMI across mid to late-life in UK Biobank.

## METHODS

### Study Setting and Participants

UK Biobank is an ongoing study of over 500,000 adults. Participants aged 40-69 years old were recruited 2006-2010 from across the UK to provide detailed information about themselves via computerized questionnaires, provide biologic samples, undergo clinical measurements, and have their health followed prospectively.^18^ In this current analyses, we excluded those with missing genetic information (n=15,221) or who were flagged as recommended for genetic analysis exclusion (n=378), and those with missing BMI (n=1,295). We also excluded participants classified as of non-European genetic ancestry (n=78,336) based on genetic ancestry principal components, because genetic predictors of AD may differ by ancestry/population stratification.^19^ After exclusions, 407,386 participants were included in the analytic sample. Ethical approval for UK Biobank was obtained from the National Health Service National Research Ethics Service and all participants provided written informed consent.

### Patient and Public Involvement

Patients were not involved in setting the research question, study design, data analysis or interpretation of the results.

### Genotyping and Genetic Risk Scores for AD

Genotyping of UK Biobank samples was conducted with two closely related arrays (Affymetrix using a bespoke BiLEVE Axiom array and Affymetrix UK Biobank Axiom array) and is described in detail elsewhere.^20,21^ Briefly, all genetic data were quality controlled and imputed by UK Biobank [downloaded on 12/1/2017] to a reference panel that merged the 1,000 Genomes Phase 3 and UK10K reference panels. A secondary imputation was completed using the Haplotype Reference Consortium (HRC) reference panel and results from the HRC imputation were preferentially used at SNPs present in both panels. Before the release of the UK Biobank genetic data a stringent QC protocol (described elsewhere) was applied at the Welcome Trust Centre for Human Genetics.^22^

To construct the AD-GRS in UK Biobank, we used summary results from the 2013 International Genomics of Alzheimer’s Project (IGAP) meta-analyzed genome-wide association study (GWAS) on late-onset AD in Caucasian populations^15^ to calculate an AD genetic risk score (GRS) for each participant. The IGAP study identified 23 loci associated with AD, including two SNPs used to characterize APOE ε4 allele status. The GRS was based on the meta-analyzed β coefficients obtained in the IGAP’s stage 1 study, which included genotyped and imputed data (7,055,881 single nucleotide polymorphisms, 1000G phase 1 alpha imputation, Build 37, Assembly Hg19) of 17,008 Alzheimer’s disease cases and 37,154 controls. We calculated the GRS by multiplying each individual’s risk allele count for each locus by the β coefficient (expressed as the log OR) for that polymorphism (Table 1) and adding the products for all 23 loci. This step weights each SNP in proportion to the observed association with AD risk (either positive or negative). The scores can be interpreted as the log OR for AD conferred by that individual’s profile on the 23 SNPs compared to a person who had the major allele at each locus. We converted the AD-GRS into a standardized z-score based on the sample mean and variance. This score has previously been shown to predict cognition and dementia-related death in the UK Biobank.^16^

**Table 1.** SNPs and Their Log Odds Ratio Estimates for the Alzheimer’s Disease Genetic Risk Score (AD-GRS)

| Marker Name | Chromosome | Closest gene | Effect Allele | Effect Estimate |
| --- | --- | --- | --- | --- |
| **AD-GRS** |  |  |  |  |
| rs6656401 | 1 | CR1 | A | 0.1567 |
| rs35349669 | 2 | INPP5D | T | 0.0663 |
| rs6733839 | 2 | BIN1 | T | 0.188 |
| rs190982 | 5 | MEF2C | G | -0.0799 |
| rs10948363 | 6 | CD2AP | G | 0.0978 |
| rs11771145 | 7 | EPHA1 | A | -0.1024 |
| rs1476679 | 7 | ZCWPW1 | C | -0.0783 |
| rs2718058 | 7 | NME8 | G | -0.0697 |
| rs28834970 | 8 | PTK2B | C | 0.0959 |
| rs9331896 | 8 | CLU | C | -0.1457 |
| rs10792832 | 11 | PICALM | A | -0.1297 |
| rs10838725 | 11 | CELF1 | C | 0.0753 |
| rs11218343 | 11 | SORL1 | C | -0.2697 |
| rs670139 | 11 | MS4A4E | T | 0.0803 |
| rs983392 | 11 | MS4A6A | G | -0.1084 |
| rs10498633 | 14 | SLC24A4-RIN3 | T | -0.1044 |
| rs17125944 | 14 | FERMT2 | C | 0.1223 |
| rs8093731 | 18 | DSG2 | T | -0.6136 |
| rs3865444 | 19 | CD33 | A | -0.0954 |
| rs4147929 | 19 | ABCA7 | A | 0.1348 |
| rs429358 | 19 | APOE | C | 1.3503 |
| rs7412 | 19 | APOE | T | -0.3871 |
| rs7274581 | 20 | CASS4 | C | -0.1390 |

### Body Mass Index

BMI was calculated based on height and weight (kg/m^2^) measured at the baseline assessment.

### Other Characteristics

Age, sex, and education were reported at baseline assessment. UK Biobank provides principal components (PCs) related to genetic population stratification, we used the first 10 PCs in our analyses to adjust for population stratification. Participants underwent a touchscreen-based cognitive battery, we focused on reaction time (i.e. simple processing speed) as a measure of cognition as it was available on the largest number of participants. Participants were timed at pressing a button as soon as two identical cards are seen on the touchscreen.

### Statistical Analysis

In preliminary analyses, we conducted analyses stratified by age at assessment (39-49 years, 50-59, and 60-73 years old). First, we evaluated the association between BMI and reaction time to confirm expected associations seen in prior research (e.g. a positive association between BMI and cognition in mid-life and a negative association between BMI and cognition in late-life). We used linear regression with reaction time as the outcome and BMI as the primary predictors with adjustment for age, age^2^, sex, and education. Next, as our primary analysis, we estimated separate age-stratified linear regressions with the AD-GRS as the primary predictor and BMI as the outcome. Models included adjustment for age, age^2^, sex, and PCs. Finally, we estimated non-linear trends for age using one linear regression model for the whole sample with AD-GRS, age, and age^2^ as primary predictors. We tested whether age trends differed by AD-GRS using interaction terms between the linear and quadratic age terms and AD-GRS. Based on this regression equation we calculated the age at which curves began to diverge. To illustrate the effects of low and high AD genetic risk we calculated predicted BMI curves based on the 10^th^ percentile (low risk) and 90^th^ percentile (high risk) of the AD-GRS. Based on these same percentiles we also estimated the earliest age at which high AD-GRS was associated with a significantly lower BMI compared to low AD-GRS. All regression models included adjustment for sex and PCs as described above. Analyses were conducted in R (version 3.3.2). All tests were two-sided with α = 0.05 and we report 95% confidence intervals (CIs).

## RESULTS

Participant baseline characteristics are shown in Table 2; 22% of the sample was aged 39-49; 32.9% aged 50-59 years, and 45.3% of the sample was 60-73 years at time of baseline assessment and 54.1% were female. The majority of participants had BMI in the normal (33%) or overweight (43%) range.

**Table 2.** Characteristics of Participants Included in the Analyses (n=407,386)

| **Participant Characteristics** | **Mean (SD) or N (%)** |
| --- | --- |
| Age, years | 56.9 (8.0) |
| 39-49 | 88,758 (21.8) |
| 50-59 | 134,163 (32.9) |
| 60-73 | 184,465 (45.3) |
| Female | 220,319 (54.1) |
| High school graduate* | 191,394 (47.5) |
| AD-GRS (z score) | 0 (1) |
| At least one APOE ε4 allele | 117,478 (28.8) |
| Reaction time, ms* | 535 (113.2) |
| BMI | 27.4 (4.7) |
| Underweight (<18.5) | 2042 (0.5) |
| Normal (18.5-24.9) | 132,315 (32.5) |
| Overweight (25-29.9) | 174,204 (42.8) |
| Obese (30-39.9) | 98,825 (24.3) |
| Morbidly Obese (40+) | 7,632 (1.9) |

\*Missing data: education: 3,701 (<1%); reaction time: 2,484 (<1%)

Age-stratified results for the association between BMI and cognition (reaction speed) are shown in Table 2. The association between BMI and cognition (reaction speed) differed by age (Table 3). For ages 39-49 years a higher BMI was associated with worse reaction time (0.13 ms slower reaction time; 95% CI: 0.01, 0.26; p=0.03). In ages 50-59 a higher BMI was associated with faster reaction time: (0.12 ms faster reaction time for ages 50-59 (95%CI: 0.23, 0.0002; p=0.045) and this effect was even stronger for ages 60-73 (0.20 ms faster reaction time 95% CI: 0.32, 0.08; p=0.001).

**Table 3.** Association between Body Mass Index and Cognition (reaction time (ms))*

| **Ages** | **N** | **$\beta$ (95% CI)** |
| --- | --- | --- |
| 39-49 | 87,973 | 0.13 (0.01, 0.26) |
| 50-59 | 132,613 | -0.12 (-0.23, -0.0002) |
| 60-73 | 180,611 | -0.20 (-0.32, -0.08) |

\*Higher values = slower reaction time; models adjusted for age, age<sup>2</sup>, sex, and education

**Table 4.** Association between AD-GRS and Body Mass Index stratified by age.*

| **Ages** | **N** | **$\beta$ (95% CI)** |
| --- | --- | --- |
| 39-49 | 88,758 | 0.00 (-0.03, 0.03) |
| 50-59 | 134,163 | -0.03 (-0.06, -0.01) |
| 60-73 | 184,465 | -0.09 (-0.12, -0.07) |

\*Adjusted for age, age<sup>2</sup>, sex, and PCs to account for confounding by population stratification

Age-stratified results for the association between BMI and the AD-GRS are shown in Table 3. In 39-49 year olds, AD-GRS was not significantly associated with BMI (0.00 kg/m^2^ per SD in AD-GRS; 95% CI: -0.03, 0.03; p=0.94). In 50-59 year olds AD-GRS was associated with lower BMI (-0.03 kg/m^2^ per 1 SD in AD-GRS; 95% CI: -0.06, -0.01; p=0.01) and this association was stronger and significant in 60-70 year olds (-0.09 kg/m^2^ per 1 SD in AD-GRS; 95% CI:-0.12, - 0.07; p<0.001).

Model-based BMI age curves showed an increasing trajectory of average BMI for individuals aged 40-50, which slowed and finally reversed so that by age 65 each additional year of age was associated with lower BMI (Figure 1). For participants with the average AD-GRS, age was associated with increased BMI (0.92 points per decade 95% CI: 0.85, 1.00) but for each decade after 40 this estimate decreased (age^2^ term) by -0.21 (95% CI: -0.06, -0.01) points. This slowing of the age-related BMI curve occurred earlier in people with high AD-GRS scores (p<0.001 for overall age by AG-GRS interactions). 1 SD AD-GRS was associated with an additional slowing of the BMI curve of -0.02 (95% CI: -0.04, 0.00) points per decade. The BMI of individuals with high versus low AD-GRS scores began to diverge after age 47 and the curves further diverged in after age 65 (Figure 1). Comparing those in the 10th to 90th percentile of AD-GRS, the BMI statistically significant lower BMI after age 56.

**Figure 1.**
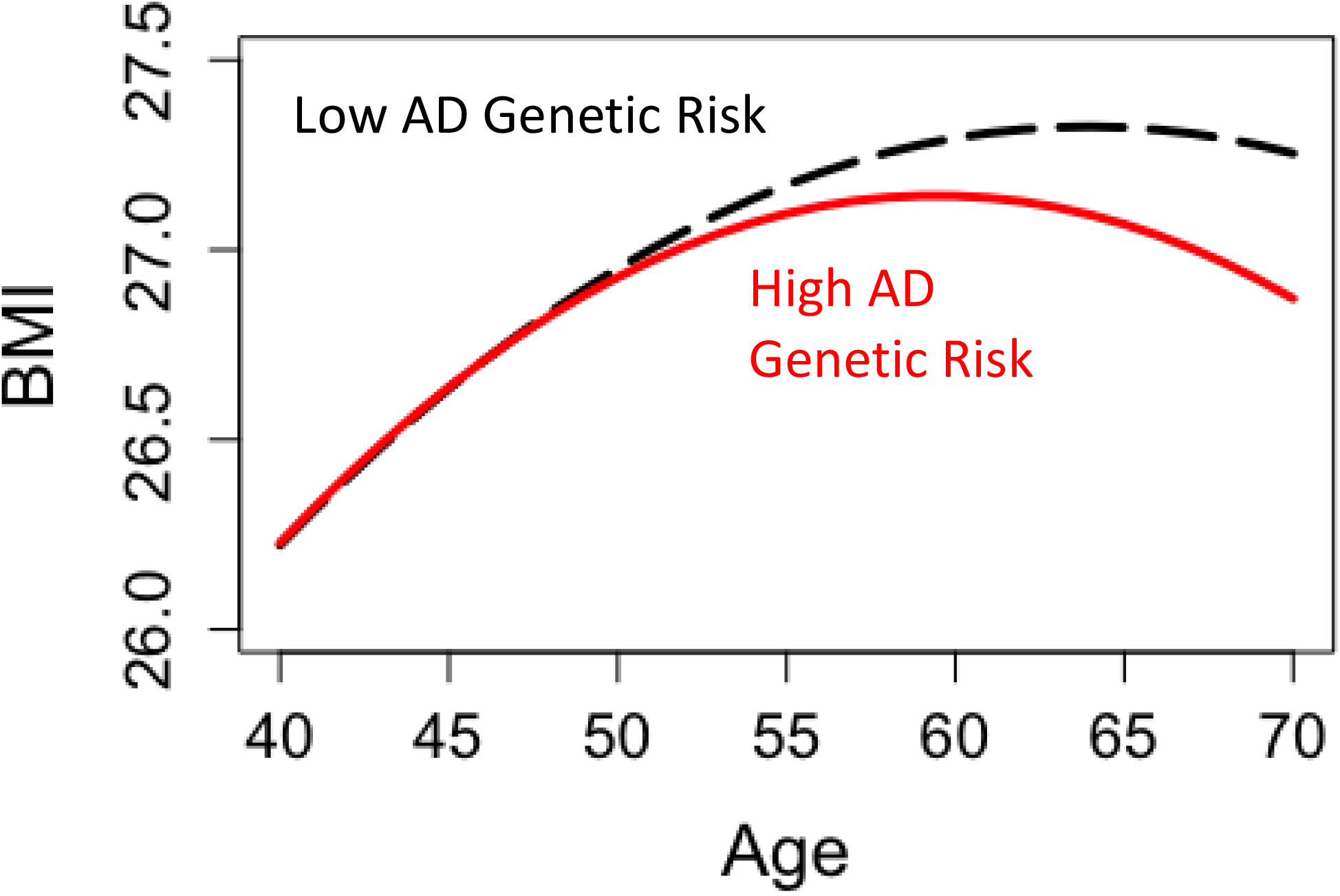
Age-related BMI curves and AD-GRS. Predicted curves for 10^th^ (low) vs. 90^th^ (high) percentile of genetic risk for AD, began to diverge at age 47 and were significantly different by age 56.

## DISCUSSION

We found evidence that higher AD-GRS predicts lower BMI in older adults. We found that the age at which BMI curves begin to diverge is in mid-life (age 47, with a statistically detectable difference by late 50s). Age-stratified analyses mirrored this finding. Likewise, a higher BMI was associated with worse cognition in mid-life and then flipped to the inverse in older ages.These findings are consistent with the hypothesis that reduced BMI or weight loss is an early manifestation of the AD process that can begin over 20 years prior to disease diagnosis.

### Evidence for BMI as an early manifestation of AD

This study expands upon prior work showing that weight loss is a predictor of subsequent AD diagnosis^4–6,12,23^ and AD neuropathologic burden.^24^ The majority of prior studies have been based on observational data in which it is not possible to establish temporal ordering of BMI relative to established markers of preclinical AD. A number of studies examining BMI in mid to late life give conflicting estimates of the age at which lower BMI is associated with AD, ranging from 20 to 8 years prior to AD diagnosis.^13,25,26^ Novel approaches can help triangulate evidence around the timing of BMI change in preclinical AD.^27^ By examining genetic risk, which is determined at birth, we provide evidence that some biological process associated with AD genetic risk affects BMI as early as age 50. Using a similar approach to our study, the Dominantly Inherited Alzheimer Network (DIAN) study assessed BMI in “preclinical AD” by comparing the point at which BMI curves began to diverge between autosomal dominant AD mutation carriers and non-carriers. They found that BMI curves for mutation carriers diverged from non-carriers as early as 17.8 years prior to expected symptom onset.^7^ Given that the average age of AD diagnosis is in the 70s we find a similarly long period (∼20 years) in which BMI may diverge between high and low AD genetic risk. Together these studies suggest processes related to AD begin many decades prior to diagnosis. We may need to target interventions in mid-life or earlier to effectively prevent AD or modify the earliest events in the AD pathologic cascade.

Our findings support the interpretation that pathophysiologic processes that culminate in AD diagnosis lead to weight loss decades earlier.^28^ Weight loss in AD may be caused by several factors, but the exact cause is unknown. Nutritional habits are worse in individuals with mild cognitive impairment or dementia;^10^ however UK Biobank participants do not have dementia and divergence in BMI curves begins years prior to symptom onset. There is also evidence that neurodegeneration and neuropathologic changes in AD are associated with lower BMI and weight loss.^9,24^ Brain regions and pathways important for metabolism, appetite, and weight maintenance such as the hypothalamic-pituitary-adrenal axis are affected in AD and may become disrupted early in the disease process.^8,29^ Our results are consistent with BMI as an early marker for changes due to AD, however, it may also be the case that pleiotropy leads to population associations between weight loss and the likelihood of AD. Although we cannot conclusively say these BMI changes are an early manifestation of AD, our findings complement prior observational studies. Additionally, even if pleiotropic pathways link the AD-GRS to weight loss, our findings confirm that lower BMI as early as midlife can predict AD and cognitive decline.

### Limitations and study strengths

There are several important caveats and limitations to our analysis. Our measurement of BMI is cross-sectional; future studies with longitudinal BMI measurements will be needed to verify age at which weight loss can begin to occur in preclinical AD. Our results are potentially influenced by selection bias due to selective survival as the AD-GRS is associated with mortality.^16^ However, this effect is likely limited because the sample is relatively young and healthy with a low mortality rate, especially due to dementia.^16^ Genetic risk for AD only explains a small percentage of variation in cognitive impairment and AD diagnosis, thus there is substantial variation in risk for AD that is not captured by our risk score. This variation reduces the ability to detect associations. These analyses were conducted in participants of European ancestry and were generally healthy and well educated; this may limit generalizability. However, there are considerable strengths to this study, particularly the large sample size, the use of measured BMI, and an innovative analytic approach. By using genetic risk, we circumvent a central challenge in interpreting prior results on weight loss, BMI and AD.

## Conclusion

Genetic factors that increase sporadic AD risk begin to predict lower BMI as early as age 47, many years prior to the average age of AD diagnosis (mid-70s). Weight loss may manifest as an early pathophysiologic change associated with AD. Additional longitudinal studies are needed to confirm the earliest detectable physiologic changes resulting from AD.

## What is already known on this topic

- Weight loss is common among Alzheimer’s disease patients in the years prior to diagnosis.
- The ages at which this association begins to manifest is unknown but has implications for prevention and treatment of dementia.

## What this study adds

- Genetic factors that increase sporadic AD risk predict lower BMI over a decade prior to the average age of AD diagnosis (mid-70s) and predicted BMI curves for low and high AD-GRS begin to diverge as early as age 47.
- These data suggest weight loss may manifest as a very early pathophysiologic change associated with AD.
- This early divergence in BMI may suggest we need to target interventions in mid-life or earlier to effectively prevent or modify the earliest events in the AD pathologic cascade.

## Data Availability

Researchers can apply to UK Biobank (www.ukbiobank.ac.uk) to access the data used in this study. No additional data is available.

## ACKNOWLEDGEMENTS

We thank UK Biobank participants and staff. The UK Biobank project is funded by the Medical Research Council, The Wellcome Trust, Department of Health for England and Wales, North West Regional Development Agency and the Scottish Executive.

## Author contributions

Willa D. Brenowitz (conception and design of the study, analysis and interpretation of data, drafting/revising manuscript for content)

Scott C. Zimmerman (analysis and interpretation of data, revising manuscript for content) Teresa J. Filshtein (analysis and interpretation of data, reviewing manuscript for content) Kristine Yaffe (interpretation of data, revising manuscript for content)

Stefan Walter (analysis and interpretation of data, revising manuscript for content)

Thomas J. Hoffmann (interpretation of data, revising manuscript for content)

Eric Jorgenson (interpretation of data, revising manuscript for content)

Rachel Whitmer (interpretation of data, reviewing manuscript for content)

M. Maria Glymour (conception and design of the study, interpretation of data, revising manuscript for content)

All authors had full access to all of the data (including statistical reports and tables) in the study and can take responsibility for the integrity of the data and the accuracy of the data analysis. The corresponding author attests that all listed authors meet authorship criteria and that no others meeting the criteria have been omitted. WDB is the guarantor.

## Funding

This work was supported by NIA grants T32-AG-049663 (WDB), K24-AG031155 (KY), RF1AG052132 (RAW and MMG), and R01AG059872 (MMG) and Community of Madrid Grant 2018-T1/BMD-11226 (SW).None of the authors’ funding organizations contributed to the design and conduct of the study; collection, management, analysis, or interpretation of the data; preparation, review, or approval of the manuscript; or decision to submit the manuscript for publication. All authors are independent from the funders.

## Competing Interests

All authors declare: no support from any organization for the submitted work; no financial relationships with any organizations that might have an interest in the submitted work in the previous three years, no other relationships or activities that could appear to have influenced the submitted work.

## Transparency declaration

The authors affirm that this manuscript is an honest, accurate, and transparent account of the study being reported; that no important aspects of the study have been omitted; and that any discrepancies from the study as planned (and, if relevant, registered) have been explained.

## Data sharing

Researchers can apply to UK Biobank (www.ukbiobank.ac.uk/) to access the data used in this study. No additional data is available.

## Ethical approval

Ethical approval for UK Biobank was obtained from the National Health Service National Research Ethics Service. All participants provided written informed consent.

## Notes

### Competing Interest Statement

The authors have declared no competing interest.

